# Systematic Living Evidence for Clinical Trials (SyLECT): a data-driven framework for drug selection in clinical trials in motor neuron disease

**DOI:** 10.1101/2025.03.09.25323612

**Authors:** Charis Wong, Alessandra Cardinali, Jing Liao, Bhuvaneish T. Selvaraj, Paul Baxter, Roderick N. Carter, James Longden, Rebecca E. Graham, Rachel S. Dakin, Suvankar Pal, Jeremy Chataway, Robert Swingler, Giles E. Hardingham, Neil Carragher, Siddharthan Chandran, Malcolm Macleod, the ReLiSyR-MND consortium

**Affiliations:** Centre for Clinical Brain Sciences, University of Edinburgh, Edinburgh, UK; Anne Rowling Regenerative Neurology Clinic, University of Edinburgh, UK; Euan MacDonald Centre for Motor Neuron Disease Research, University of Edinburgh, Edinburgh, UK; Medical Research Council Clinical Trials Unit at UCL, Institute of Clinical Trials and Methodology, University College London, London, UK; UK Dementia Research Institute at University of Edinburgh, University of Edinburgh, Edinburgh bioQuarter, Chancellor’s Building, 49 Little France Crescent, Edinburgh, UK; Queen Square Multiple Sclerosis Centre, Department of Neuroinflammation, UCL Queen Square Institute of Neurology, Faculty of Brain Sciences, University College London, London, UK; National Institute for Health Research, University College London Hospitals, Biomedical Research Centre, London, UK; Centre for Discovery Brain Sciences, University of Edinburgh, Edinburgh, UK; MRC Institute of Genetics & Cancer, The University of Edinburgh, Edinburgh, UK

**Keywords:** Motor neuron disease, neurodegenerative diseases, evidence synthesis, clinical trials, drug repurposing

## Abstract

Despite many promising preclinical studies and decades of clinical trials, there remains a paucity of effective disease-modifying drugs in motor neuron disease. We aimed to develop a systematic and structured data-driven framework to identify, evaluate and prioritise candidate drugs for clinical trials, specifically for the Motor Neuron Disease-Systematic Multi-Arm Adaptive Randomised Trial (MND-SMART; NCT040302870). We developed the Systematic Living Evidence for Clinical Trials (SyLECT) platform as a modular framework integrating emerging data from different domains to inform prioritisation of candidate drugs. Current domains incorporated include published clinical, animal *in* vivo, and *in vitro* literature; in house *in vitro* high throughput drug screening; pathway and network analysis; and pharmacological, feasibility and clinical trial data from drug, chemical, and clinical trial databases. In this approach, we first identify a list of candidate drugs from these domains then select drugs for further consideration based on drug properties, feasibility, and expert opinion. For prioritised drugs we then generate, evaluate, and synthesise further evidence from across data domains. Using automated workflows and interactive web applications, we produce snapshot “living evidence summaries” to inform expert panel decisions on prioritisation of candidate drugs for MND-SMART. The third drug selected for MND-SMART and the first using this framework is amantadine. We demonstrated the feasibility of a systematic data-driven framework to inform prioritisation of candidate drugs for clinical trials in motor neuron disease, with potential for wider application across diseases where there is unmet clinical need.

**Key messages:** *What is already known on this topic:*

- Despite extensive preclinical research and clinical trials for disease-modifying treatments in motor neuron disease, translational success remains elusive.
- Advances in research across biological domains presents a wealth of data to guide prioritisation of candidate drugs for clinical trials.

*What this study adds:*

- This study demonstrates the feasibility of using a systematic, modular, data-driven framework to inform prioritisation of candidate drugs for an adaptive platform trial in motor neuron disease.

*How this study might affect research, practice or policy:*

- The framework could be applied to inform prioritisation of drugs for clinical trials in other diseases, especially adaptive platform trials in neurodegenerative diseases.

## Introduction

Motor neuron disease (MND, alternatively amyotrophic lateral sclerosis (ALS)), is a progressive, disabling and ultimately fatal neurodegenerative disease. In many countries including the United Kingdom (UK), riluzole remains the only approved disease-modifying drug, with an average survival benefit of 2-3 months.(1) Edaravone, masitinib and AMX0035 have emerged as promising candidates with positive trials in highly stratified cohorts.(2–5) However, evidence of generalisable survival benefit remains limited, and none of these drugs have been granted marketing authorisation for MND/ALS in Europe. The paucity of effective disease-modifying treatments despite numerous clinical trials and promising preclinical studies reflects the challenges in selecting and evaluating drugs for clinical trials, compounded by limitations in our understanding of MND disease biology.(6) However, recent adaptive platform trials such as the Motor Neuron Disease – Systematic Multi-Arm Adaptive Randomised Trial (MND-SMART; EudraCT Number: 2019-000099-41; ClinicalTrials.gov registration number: NCT04302870),(7) HEALEY ALS Platform Trial (ClinicalTrials.gov registration number: NCT04297683),(8) and the TRICALS ‘MAGNET’ platform trial (EudraCT number: 2020-000579-19)(9) provide efficient infrastructures to evaluate pipelines of candidate drugs, offering significant reduction in overall sample size, cost and time compared to consecutive two-arm trials with traditional fixed designs.(6, 10)

There is, therefore, an urgent need to optimise drug selection and prioritisation for clinical trials in MND. Historically, these decisions have been largely informed by mouse studies, with inherent limitations in modelling human disease. Most of these studies use superoxide dismutase (SOD1) mouse models which do not recapitulate the TAR DNA binding protein 43 (TDP-43) proteinopathy seen in 97% of people with sporadic MND.(11) A reported lack of reproducibility and rigour of many of these studies likely further contributes to translational failure.(12)

These limitations might be mitigated by taking a more comprehensive, unbiased and systematic approach to selection and prioritisation of drugs for clinical trials. Structured expert led guidance have been used in in secondary progressive multiple sclerosis,(13) Alzheimer’s disease,(14) and Parkinson’s disease,(15) where expert committees are convened to suggest relevant mechanisms and longlist drugs acting on these mechanisms. Drug “Curriculum Vitaes” (CVs) with a standard template including information on pharmacodynamics, pharmacokinetics, mechanism of action, and *in vitro*, *in vivo* and clinical trials evidence base are completed by members of the committee and scored to inform further rounds of shortlisting. However, without systematic evidence synthesis, incomplete capture of information to this process may result in promising candidate drugs being missed and evidence being overstated, while inconsistencies in compilation and presentation of information by contributors from different backgrounds may introduce bias.(13)

We used clinical and preclinical systematic reviews to guide expert panel discussions for drug selection in the Multiple Sclerosis – Secondary Progressive Multi-Arm Randomisation Trial)(16, 17) and for the first two treatment arms of MND-SMART.(7, 18) This phase III multi-arm multi-stage trial began recruiting in February 2020, with three treatment groups comprising placebo, memantine, and trazodone, and by October 2022 had randomised 400 participants. The trial design enables discontinuation of treatment arms that are ineffective and addition of new treatment arms. The trial investigators seek to identify and prioritise a pipeline of suitable, ‘trial ready’ candidate drugs for future addition. The main challenge is in producing an up-to-date summary of the available evidence, given the time taken to complete systematic reviews. Further, an approach based purely on the published literature may overlook other therapeutic approaches, which might be suggested for instance by unbiased drug screening assays or disease-specific pathophysiology (to the extent that this is known).

Recent innovations in systematic review and evidence synthesis technology, including automation tools, machine-learning (ML) and text-mining techniques, have made it feasible to perform living systematic reviews at scale, and to produce living evidence summaries to inform practice and policy.(19, 20) In parallel, advances in human-induced pluripotent stem cells, gene editing and multi-omics technology have facilitated rapid expansion of our understanding of MND disease and related biology and the identification of putative therapeutic targets and pathways, some of which are shared across neurodegenerative diseases.(21) Automated high throughput phenotypic *in vitro* drug screening technology has enabled comprehensive screening of compound libraries representing approved drugs and potential clinical candidates on human-derived models at scale and at speed.(22) The past decade has also seen rapid development and progress in computational and bioinformatics tools and resources which integrate and leverage multiple data types to generate drug-target-phenotypic hypotheses. Noting these advances across disciplines, we aimed to develop a systematic and structured framework to integrate data from different domains (including expert opinion) to identify, evaluate and prioritise candidate drugs for evaluation in MND-SMART.

## Results

Our drug identification, evaluation, and prioritisation process for the next arm of MND-SMART is summarised in Figure 1. In September 2019, 20383 publications were retrieved from our Repurposing Living Systematic Review – Motor Neuron Disease (ReLiSyR-MND) living clinical search, of which 6138 publications describing 798 interventions were included. Of these, 196 interventions described in 1660 clinical publications met the ReLiSyR logic, and of these, 45 drugs described in 1082 clinical publications met the longlisting criteria in place at that time (listed in the British National Formulary, prescription-only medicine available in oral formulation and not previously in the 2017 ‘red list’ during selection of the first two arms of MND-SMART, deemed by investigators to be appropriate for MND-SMART). For these publications we undertook annotation and data extraction from clinical, animal *in vivo* and *in vitro* publications within ReLiSyR-MND.

**Figure 1.**
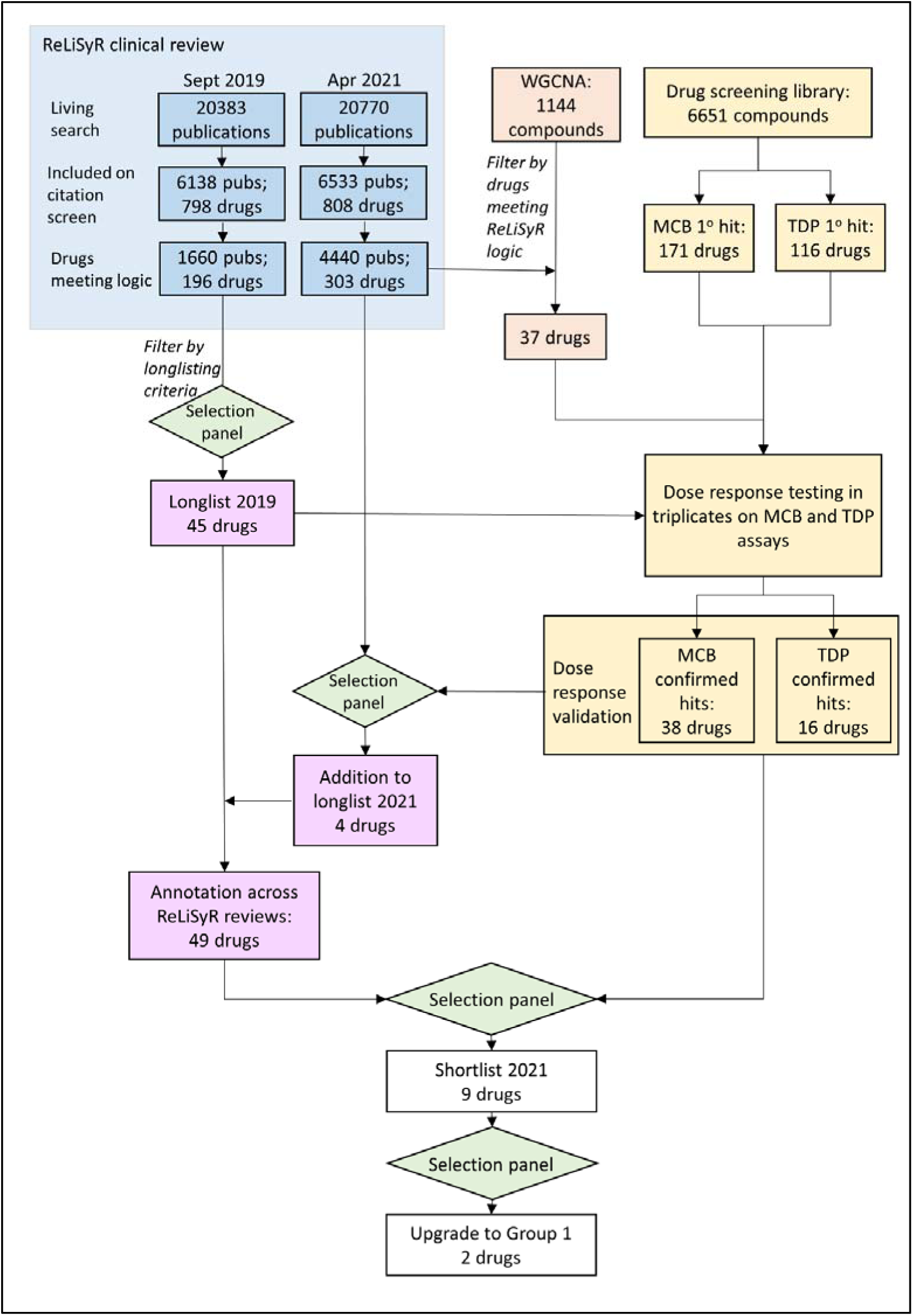
Identification, evaluation and prioritisation of candidate drugs. MCB: monochlorobimane; pubs: publications; WGCNA weighted gene co-expression network analysis.

In April 2021, the number of clinical publications retrieved in our living search had increased to 20770, of which 6533 publications describing 808 interventions were included. 303 interventions described in 4440 publications met the ReLiSyR logic. In parallel, on primary *in vitro* screening, we identified 171 positive astrocyte monochlorobimane (MCB) hits and 116 positive hits which reduced wild-type and/or mutant TDP-43 protein aggregates in HEK293 cells and human stem cell-derived motor neurons under oxidative stress (sodium arsenite). We identified 1144 compounds of interest by weighted gene co-expression network analysis (WGCNA) using TargetALS RNA-sequencing data. We performed dose response *in vitro* screening in triplicates on astrocyte MCB and motor neuron TDP-43 assays for all drugs longlisted in 2019, all positive hits on primary screen and 37 compounds identified on WGCNA which met ReLiSyR logic. 38 astrocyte MCB hits and 16 TDP-43 hits were confirmed on dose response retest. We added four drugs meeting ReLiSyR logic and showing activity on *in vitro* screening to our longlist and generated further evidence across all domains for longlisted drugs.

In October 2021, we reviewed the above data, and mechanistic data from DrugBank (www.drugbank.ca) and PubChem with a particular focus on (i) drugs with high ReLiSyR scores in the current review and in 2017, (ii) ‘rapid risers’ – drugs with large improvements in ReLiSyR rankings from 2017 and (iii) drugs featuring in the current review of ReLiSyR which were not included in the 2017 review, and (iv) drugs showing compelling evidence from other sources including published preclinical studies, *in vitro* screening, pathway and network analysis, and preprints. The group shortlisted nine drugs for further evaluation. Following expert panel review of the expanded literature search, scoping of manufacturing feasibility and multiparametric phenotypic screening in triplicate with dose response testing, two drugs, amantadine and ropinirole were upgraded to Group 1 in January 2022, with amantadine selected as the third experimental arm for MND-SMART. ReLiSyR clinical data used for the shortlisting process are shown in Figure 2.

**Figure 2.**
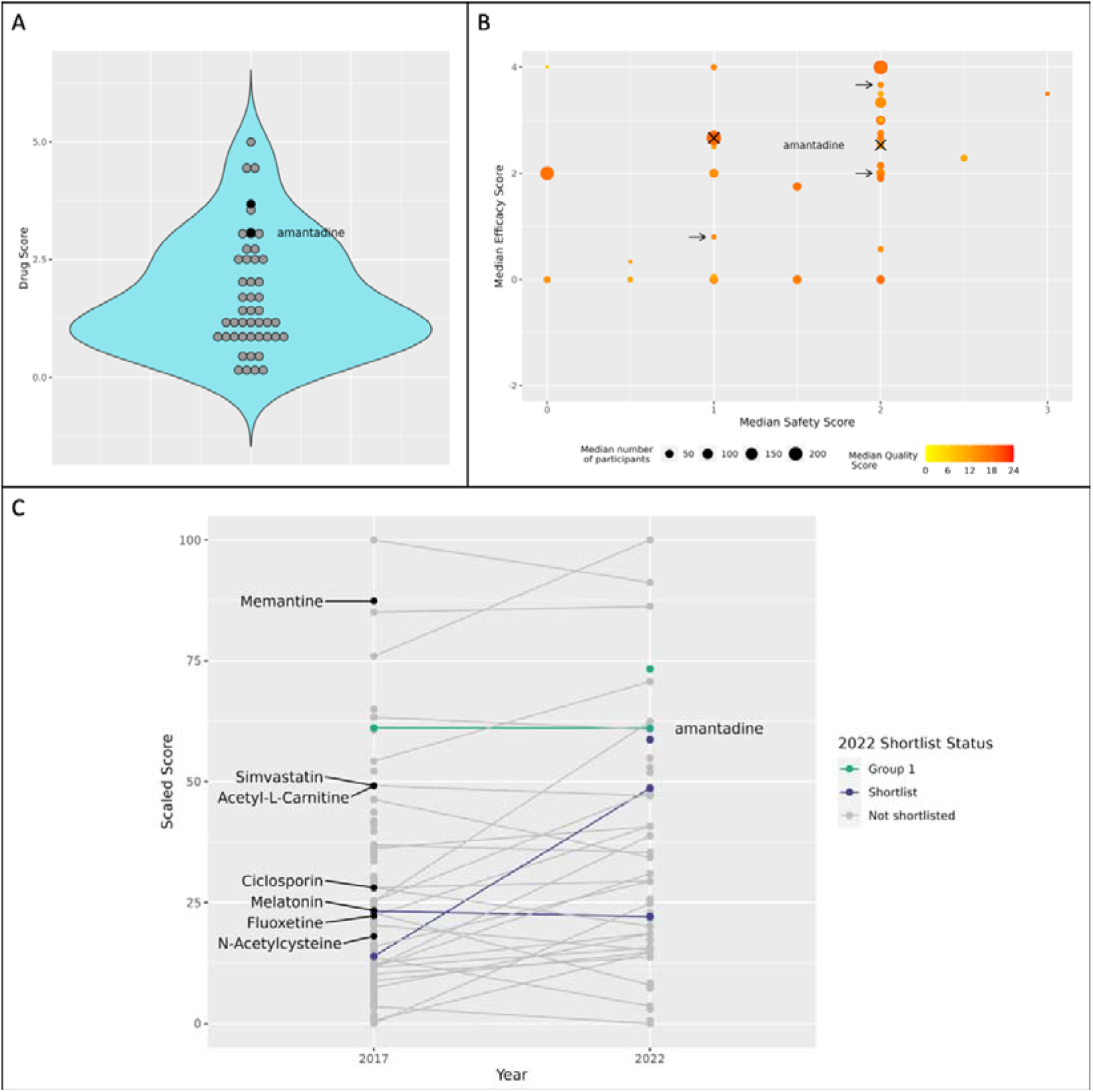
Scores from ReLiSyR-MND clinical review used to inform prioritisation of candidate drugs (as of 28 April 2022). (A) Violin plot showing distribution of ReLiSyR drug score for longlisted drugs. Group 1 drugs are shaded in black. (B) Bubble plot showing ReLiSyR clinical review subscores for all longlisted drugs. Drugs recommended for clinical trial are marked with crosses. Other shortlisted drugs are marked with arrows. (C) Changes in drug scores for drugs between previous review concluded in 2017 and the current review. Shortlisted drugs are labelled. Scores shown are rescaled by min-max normalisation, where 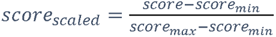

### Amantadine

Amantadine is a synthetic tricyclic amine with antiviral, antiparkinsonian and antihyperalgesic activities. Amantadine is licensed in the UK for treatment of Parkinson’s disease and post-herpetic neuralgia, and treatment and prophylaxis of influenza A. Amantadine is also used for fatigue in multiple sclerosis but is not licensed for this indication. Amantadine is a weak dopamine agonist (which may give rise to its antiparkinsonian effects) and interferes with a viral protein, M2. Amantadine also targets NMDA glutamate receptors, and this may be relevant to glutamate-induced neurotoxicity in neurodegenerative diseases.

We identified 59 publications describing the use of amantadine in a total of 2201 participants (Table 1), along with 2 preprints in multiple sclerosis (171 participants) as of 28 April 2022. Amantadine ranked 7 out of 47 longlisted drugs. Efficacy (median 2.53, range: −2 - 4) and safety (median 2; range: 0-3) subscores were favourable. Amantadine has previously been trialled in ALS in 1981 in a small, double-blind crossover randomised controlled trial of amantadine (100 mg three times daily), guanidine and placebo over 6-month treatment periods.(34) While the study did not show significant improvement in a measure based on functional performance and strength, no quantitative data were presented. Amantadine was tolerated well.

**Table 1.** Characteristics and number of patients / participants of clinical studies for amantadine identified via ReLiSyR-MND.

| **Disease** | **Study Type** | **Number of Studies** | **Number of Participants** |
| --- | --- | --- | --- |
| Huntington's disease | Interventional | 3 | 57 |
| Motor neuron disease | Interventional | 1 | 10 |
| Multiple sclerosis | Interventional | 11 | 452 |
| Parkinson's disease | Interventional | 34 | 1240 |
|  | Observational | 10 | 442 |
| **Total** |  | **59** | **2201** |

Intracellular TDP-43 aggregates are seen in 97% of MND post mortem cases and in 40% of frontotemporal dementia (FTD) post mortems, and it is likely to contribute to neuronal death.(35) Thus, drugs able to reverse the pathological accumulation of TDP-43 aggregates are strong candidates for testing in clinical trials. On *in vitro* screening, amantadine was effective in clearing TDP-43 cytoplasmic aggregates *in vitro* in human neurons derived from people with MND, significantly reducing the number and size of TDP-43+ aggregates (Figure 3).

**Figure 3.**
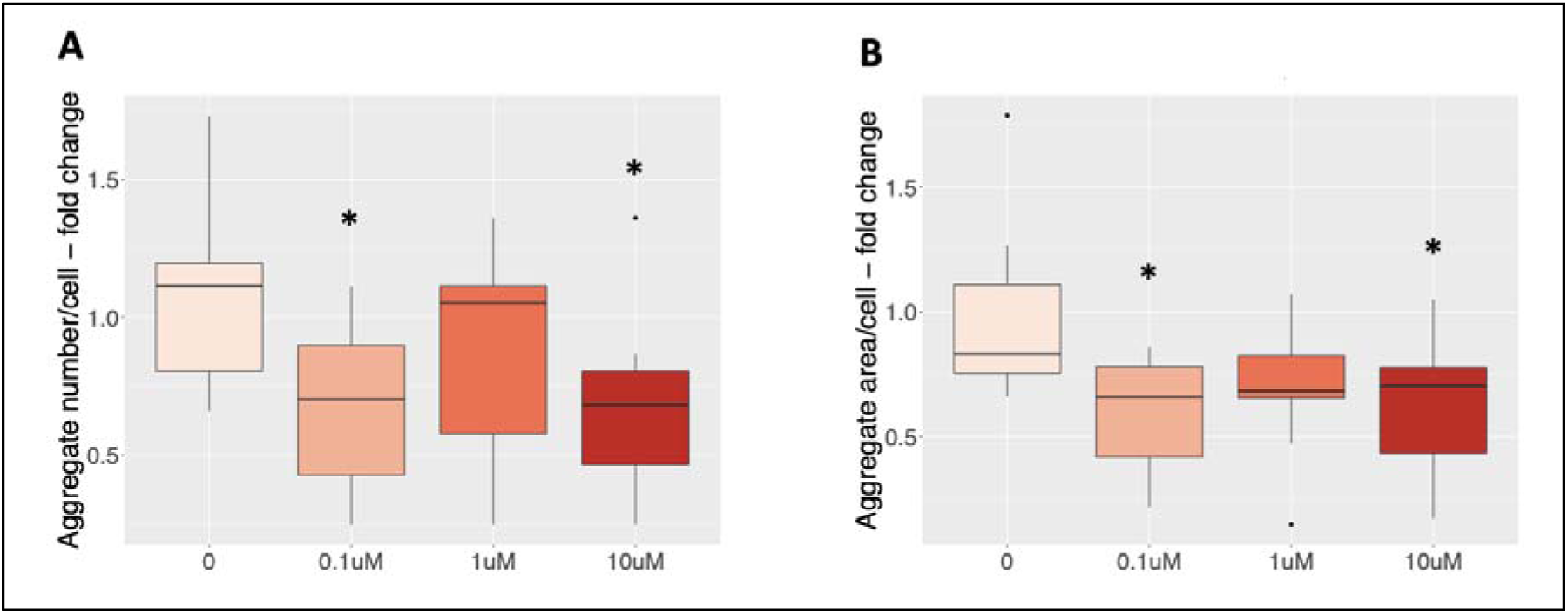
Dose-response curve for treatment of human stem cell-derived spinal motor neurons with amantadine depicting reduction in number (A) and area (B) of TDP-43+ aggregates. *p < 0.05, two-sample Student’s t-test to vehicle control (0uM).

In parallel, using target prediction by SEA and STRING network analysis, we built a predicted protein-protein interaction network for amantadine (Figure 4). There was a strong overlap between proteins within this network and association with MND based on disease association scores from the Open Targets Platform.

**Figure 4.**
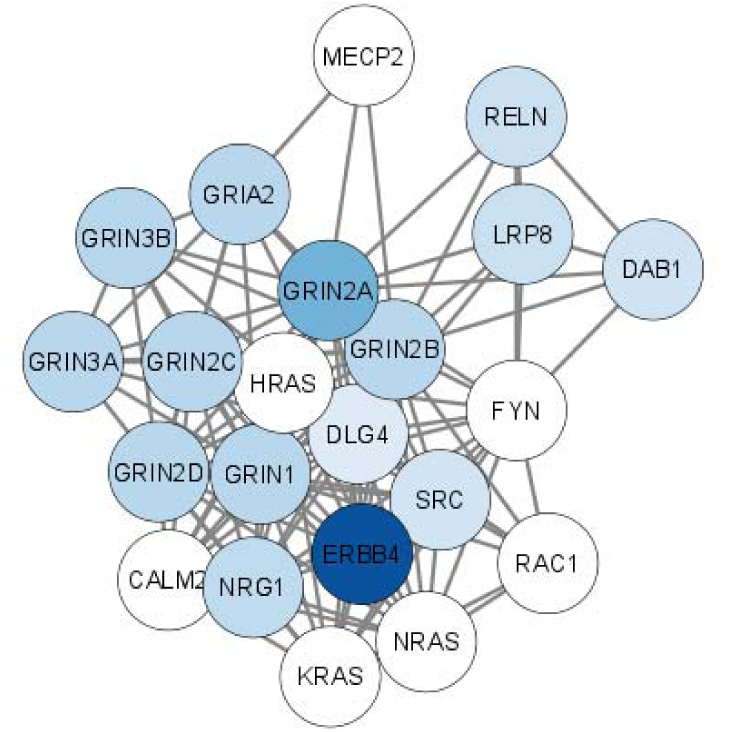
Protein-protein interaction network for amantadine based on target prediction using Similarity Ensemble Approach and expansion by STRING database to the nearest 20 proteins. Shade of nodes represent MND disease association scores for targets obtained from Open Targets Platform, with darker shades representing stronger association.

Amantadine is well absorbed orally, has good predicted BBB penetrance and can be given as a once daily oral solution, matching the other investigational medicinal products currently tested within MND-SMART.

## Discussion

Translational success has remained elusive across neurodegenerative diseases. However, the accelerating pace of research and advances across biological domains: ‘-omics’ fields from genomics, metabolomics, proteomics, transcriptomics to epidemiology with wider and more in-depth phenotypic capture of population data via disease registries, and wearable technologies provide an ever-expanding wealth of data which can be instrumental in achieving improved clinical outcomes. These data may indeed hold key insights towards improving our understanding of underlying disease mechanisms and developing effective treatments to slow, stop and reverse these processes. The emergence of adaptive platform trials in neurodegenerative diseases brings a new challenge – to identify, evaluate, and select the best available candidate drugs for evaluation in clinical trials within the timeframe of trial adaptation epochs. To unlock the potential of large and rapidly expanding amounts of data which are often siloed to inform such decisions, developing methods to provide a continually up-to-date, systematic, broad-scope synthesis and mapping of emerging evidence across relevant domains is crucial.

As a starting point, we built upon our previous clinical and preclinical systematic reviews informing the selection of the first two arms of MND-SMART. To make it feasible for ReLiSyR to provide up-to-date synthesis of the rapidly expanding literature, our updated methodology incorporates living updates of systematic searches and automated removal of duplicate records, machine learning-assisted citation screening for inclusion, automated annotation of drug and disease mechanisms studied in each publication. Further updates to the Systematic Review Facility (SyRF) platform(19, 24, 36) enables efficient project management, data management, implementation of systematic review tools and crowdsourcing of annotations, efficient workflows via R and R Shiny to automate scoring, analysis, reporting including visualisation and updates to MND-SOLES-CT.(19, 25, 37)

Our approach to drug selection has evolved with accumulating experience and expansion of our networks to incorporate more domains and gather more in-depth data with adaptations to better suit current clinical trial protocol and requirements. We adopted a modular approach to enable flexibility to incorporate other suitable data domains. High-throughput *in vitro* screening of comprehensive drug libraries allows evaluation of drug-target hypotheses in the context of all existing drug-target classes to facilitate benchmarking and prioritization. Further integration with pathway and network analysis provide means of evaluating disease-specific pathophysiology for candidate drugs identified via ReLiSyR. The addition of other domains has had a synergistic effect on our evidence synthesis, with each domain informing further development of other domains – for example, where we have used ReLiSyR data to select drugs for more in-depth *in vitro* screening and vice-versa.

As our primary aim is to inform drug selection within trial adaptation cycles for a phase III multi-arm multi-stage trial, drug repurposing – using an established drug in a novel indication – offers significant advantages over evaluation of novel compounds with shorter development timelines, availability of safety data and lower costs, and is thus our current focus. Novel compounds of interest identified can be evaluated further in preclinical studies and early phase clinical trials to gather further data for future consideration in MND-SMART.

### Limitations

One of our challenges is to evaluate, synthesise and summarise large amounts of data from different domains in an intuitive, informative, and understandable manner for our expert panel. Currently, for longlisted drugs, we do this for each domain separately and present these in our drug CVs and via our app. With increasing numbers of drugs to consider, a scoring system which produces one overall score reflecting the suitability of each drug for evaluation in clinical trials using evidence across all domains could prove useful. To achieve this, we need to develop methods to evaluate the strength and relevance of each domain in comparison to the others and weigh them accordingly. However, this is challenging in a disease where few effective disease-modifying treatments exist and the ‘absolute truth’ of how informative each of these domains are in predicting the likelihood of translational success remains unknown.

The other challenge is to produce accurate ‘real-time’ evidence summaries. The innovations in ReLiSyR have enabled us to produce a ‘real-time’ living evidence summary of publications retrieved from biomedical databases, deduplicated, screened for inclusion and annotated with drug and disease studied to inform the longlisting of candidate drugs. However, more in-depth annotation of publications for longlisted drugs is currently a time-consuming process fully dependent on human reviewers. We have thus limited the number of longlisted drugs for in-depth annotations within ReLiSyR to ensure these are completed within trial adaptation timelines, noting a lead time required for trial protocol amendments, discussions with regulators and sponsors, and manufacturing prior to inclusion of the selected drug in MND-SMART.

The current MND-SMART master protocol employs the same eligibility criteria, dosing regimen and route across all treatment arms, with all investigational medicinal products manufactured as matched oral solutions, thus limiting the scope of drugs which can be feasibly evaluated within the trial. Planning of a protocol amendment to allow different eligibility criteria, dosing regimens and routes for different treatment arms is ongoing.

### Future directions

Other suitable domains, such as epidemiology, clinical trial data, and other multiomics data can be added to our modular framework to increase the breadth of evidence synthesised.

Ongoing progress in other systematic review, evidence synthesis and automation tools including those in other Systematic Online Living Evidence Summary projects are potentially transferrable to our workflow.(19, 20) The expanding datasets from human annotations and data extraction in ReLiSyR can be used to train and validate further machine learning models to further improve efficiency. Developing evidence synthesis at the levels of pathways and targets in addition to current synthesis at the level of drugs may be helpful to inform prioritisation of targets and pathways for further research and drug development, and for consideration of combination treatments. The overall framework could be adapted and implemented in other diseases, especially in adaptive platform trials.

## Conclusion

We demonstrate the feasibility of using a modular framework with data from different domains to inform systematic prioritisation and evaluation of candidate drugs for an adaptive platform trial in MND, with potential applications in other diseases. Work to expand the breadth, depth and efficiency of evidence synthesis within the framework is ongoing.

## Material and methods

The principle underpinning our work is that prioritisation of candidate drugs for an ongoing adaptive platform trial is best informed by the systematic gathering, evaluation and synthesis of all evidence available at that time. To best inform drug selection at the pace required for trial adaptations, this process must be able to rapidly and efficiently identify then prioritise candidate drugs, followed by more in-depth evidence generation, evaluation and synthesis for prioritised drugs. This expansion in scope, breadth and depth of the evidence synthesised, adaptation to best suit clinical trial requirements, and improvement in efficiency of evidence synthesis over time also needs to proceed at pace. Against this background, we developed the SyLECT (Systematic Living Evidence for Clinical Trials) Framework (Figure 5). Importantly, the automation tools and algorithms used to synthesise information from different domains are subject to continual improvement; the current implementation of SyLECT is described below.

**Figure 5.**
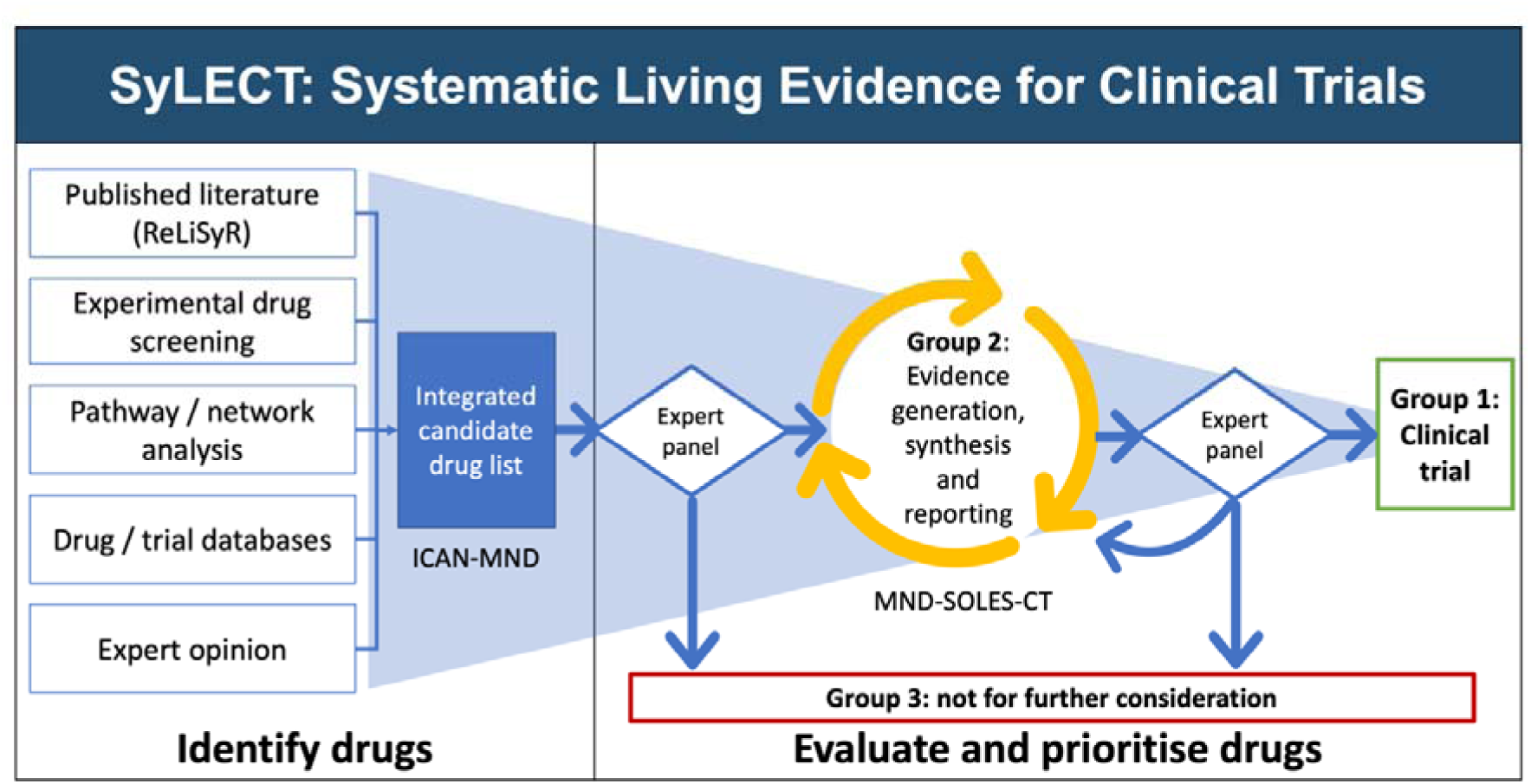
The Systematic Living Evidence for Clinical Trial (SyLECT) framework.

We identify suitable and available domains of data to provide evidence on efficacy, safety, feasibility, and pharmacological and biological profiles of candidate drugs. For the current iteration we are again seeking an existing drug suitable for repurposing. We began with summarising published data for drugs tested in neurodegenerative diseases (curated in the ReLiSyR-MND platform (23); stable version: https://camarades.shinyapps.io/relisyr-v1/) and recommendations from experts. We have since added consideration of domains describing findings from *in vitro* high throughput phenotypic drug screening, pathway and network analysis, and mining of drug, compound, and clinical trial databases. We generate an integrated candidate drug list using these domains and filter drugs with our prioritisation criteria via the ICAN-MND (Integrated Candidate Drug List for MND) app, an interactive web application (demo version: https://camarades.shinyapps.io/ican-mnd-demo). Our expert panel create a longlist of drugs from the filtered list. For longlisted drugs, we generate, evaluate, and synthesise further evidence across domains. We report living evidence summaries using MND-SOLES-CT (Motor Neuron Disease – Systematic Online Living Evidence Summary for Clinical Trials; demo version: https://camarades.shinyapps.io/mnd-soles-ct-demo), an interactive web application which can be explored by the expert panel to inform their decisions on drug shortlisting and selection at trial adaptation boundaries.

### Identifying candidate drugs

#### (i) Repurposing Living Systematic Review – Motor Neuron Disease (ReLiSyR-MND)

ReLiSyR-MND is a ML-assisted, three-part living systematic review of clinical studies of MND and other neurodegenerative diseases (Alzheimer dementia, FTD, Huntington disease, Parkinson disease and multiple sclerosis) which may share similar pathways; animal *in vivo* studies of MND and FTD models (the latter included because of the pathological overlap between FTD and MND); and *in vitro* literature describing human induced pluripotent stem cell based MND and FTD models. ReLiSyR builds upon our clinical and preclinical systematic reviews which concluded in 2017 informing the first two arms of MND-SMART. (7, 18) The protocol of ReLiSyR-MND has been updated with accumulating experience since its conception. The complete record of the protocol, including changes made, is available on Open Science Framework.(23) Using Systematic Review Facility (SyRF; syrf.org.uk; RRID:SCR_018907),(24) a web application for systematic reviews, we conduct a living search with weekly automated retrieval of citations from Pubmed via an Application Programming Interface (API). Using a ML-algorithm trained on more than 5000 dual-screened human decisions (sensitivity 95%, specificity 81%, precision 67%), citations are screened for inclusion based on title and abstract. We use text mining (Regular Expressions deployed in R(25)) to annotate included publications for drug and disease studied. We then select drugs described in at least one clinical publication in MND OR in clinical publications in two other diseases of interest (the “ReLiSyR logic”).

#### (ii) *In vitro* high throughput drug screening

We performed multi-parametric high throughput screening of the Prestwick Chemical Library, the TargetMol drug repurposing library, and the TargetMol Natural Product Library (6651 compounds) spanning all major drug-target classes, including United States Food and Drugs Administration-approved drugs using validated in-house phenotypic assays. Current assays include human astrocyte antioxidant assays and a TDP43 protein aggregation assay. Drug screening methods are detailed in a separate publication (in progress).

Primary screening for these assays were performed in 384-well format to industry standards of reproducibility and signal-to-noise using liquid handling robotics and automated image acquisition and analysis (ImageXpress-confocal platform), with libraries professionally stored, curated and plated by BioAscent.

#### (iii) Pathway and network analysis

We perform weighted gene co-expression network analysis (WGCNA) on RNA-sequencing (RNA-seq) data from the TargetALS consortium of postmortem brains of people with MND and controls.(26) From this we identify network modules significantly associated with MND (because these may signify possible underlying dysfunction or secondary pathophysiological effects) and use this to identify compounds which target proteins within these networks. Where these targets are not amenable to drug treatment in humans due to toxicity, feasibility, or pharmacological issues such as poor BBB penetrance of existing drugs, we conduct further analysis to identify potential upstream or downstream targets and candidate drugs which may act on those targets.

#### (iv) Mining drug, compound and trial databases

We mine ChEMBL,(27, 28) admetSAR2.0,(29) and ClinicalTrials.gov to obtain data on pharmacology, chemical and physical properties, mechanisms of action, predictions on BBB penetrance, and MND clinical trials.

#### (v) Expert panel

We gather recommendations from our panel consisting of MND triallists, clinicians, scientists, and experts in drug screening, pharmacology, and systematic reviews.

Reporting candidate drugs identified

We report all candidate drugs identified across the domains using ICAN-MND. In this app, users can select categories of evidence of interest (protein aggregation screening hits, astrocyte oxidation assay hits, drugs in any ReLiSyR included publications, drugs meeting ReLiSyR drug/disease logic, current longlisted drugs and/or WGCNA predictions) and filters (admetSAR2.0(29) prediction of BBB permeability, oral formulation, prescription only medicine, listed in the British National Formulary). The app generates an Euler plot visualising number of drugs in each set, and where these sets overlap, and tabulate drugs meeting the criteria selected.

#### Evaluating and prioritising candidate drugs

Using our framework, we inform expert panel decisions in prioritising candidate drugs into three groups: Group 1: suitable for evaluation in clinical trial; Group 2: for further evidence generation, synthesis, and reporting; and Group 3: not for further consideration (Box 1).

##### Box 1

**Priority groups for candidate drugs**

Group 1: Recommended for clinical trial

Group 2: Recommended for further evidence generation, synthesis, and reporting.

- Group 2A: Drugs suitable for imminent evaluation in MND-SMART, pending further evidence synthesis and evaluation.
- Group 2B: Drugs not suitable for imminent evaluation in MND-SMART but may yield important data for pathway and network analysis or be suitable for other clinical trials including early phase trials.
- Group 2C: Drugs not suitable for imminent evaluation in MND-SMART, due to pharmacological and feasibility profiles. Candidates for further mechanistic and drug development studies.

Group 3: Not recommended for further evaluation.

We identify candidate drugs meeting ReLiSyR logic, or those with positive hits on primary screening in relevant *in vitro* screening assays, or promising WGCNA predictions from TargetALS RNA-Seq data,(26) or suggestions from the expert panel. From these, we take forward candidates meeting all the following criteria: listed in the British National Formulary,(30) status as prescription only medicine (to prevent self-administration of over-the-counter trial drugs which may compromise trial integrity), availability in oral formulation, and predicted BBB permeability. We remove candidates which have been previously deemed unsuitable for clinical trials by the expert panel (due to prior definitive testing in MND clinical trials, lack of biological plausibility or poor safety profile) unless there is overwhelming new evidence to justify reconsideration. The expert panel reviews reported safety, biological plausibility, feasibility, and suitability of remaining candidate drugs to create a longlist for further evaluation (Group 2A). The expert panel may assign candidate drugs which do not meet the criteria above to lower priority groups (Group 2B or 2C) for further evaluation.

For longlisted drugs, we curate further evidence across the different domains:

(i) ReLiSyR-MND: We recruit and train a crowd of reviewers to annotate and extract outcome data from publications for longlisted drugs, using the SyRF platform.(24) Each publication is annotated independently by two reviewers, with discrepancies reconciled by a third reviewer. For the clinical review, using a predefined metric, we programmatically score each drug in each publication on a combination of efficacy and safety and weighted by study quality scored using a modified GRADE (Grading of Recommendations, Assessment, Development and Evaluations) framework and study size.(23) We combine data for each drug across publications taking the median publication score for each drug and number of contributing publications, and rank them accordingly. For *in vivo* and *in vitro* studies, we generate forest plots summarising the treatment effect of each longlisted intervention. Where there are three or more publications reporting the effect of an intervention in at least five independent experiments we conduct a formal meta-analysis.
(ii) Experimental drug screening: To validate positive hits from primary in vitro screens we perform 8-point semi-log dose response confirmation experiments, using alternative suppliers. We performed secondary screening protocols using protein aggregation assays and neurotox assays. Details of these are within a separate publication by the drug screening team.
(iii) Pathway and network analysis: We perform protein-target prediction for each longlisted drug using the structural similarity ligand-based target prediction approach, SEA (Similarity Ensemble Approach)(31), using Z score >1.96 and MaxTc (Tanimoto coefficient) greater than 0.4 as thresholds) with compound SMILES (simplified molecular-input line-entry system) as input. We then build protein-protein interaction networks using STRING database by expanding from predicted targets to the nearest 20 proteins.(32) We then perform pathway enrichment analysis to identify biological pathways hit by each compound. Separately, we search the Open Targets Platform for targets associated with “amyotrophic lateral sclerosis” and their disease association scores based on genetic associations, somatic associations, drugs, pathways and system biology, RNA expression and animal models. We map predicted protein-protein interaction network for each drug to these disease association scores.
(iv) Drug databases: We mine data from the ChEMBL(27, 28) database to gather pharmacological information such as pharmacokinetics and pharmacodynamics data, chemical and physical properties, and mechanism of action. We gather data on prescribing, safety, licensing, and medicinal forms from the British National Formulary.(30)

We provide a curated living evidence summary from all the domains using an interactive application, MND-SOLES-CT and use R Markdown (33) to automate generation of timestamped parameterised drug CVs for drug selection to inform expert panel discussions. The expert panel meets at trial adaptation boundaries to discuss candidates for shortlisting. For shortlisted drugs, we expand our literature search to include preprint servers, and extract further data on safety, tolerability, and dosing. We evaluate suitability of shortlisted drugs based on current trial protocol and consider impact on trial eligibility criteria, dosing schedule and feasibility in manufacturing compounds in forms matching other current investigational medicinal products within MND-SMART. Candidate drugs recommended for clinical trials are re-assigned from Group 2A to Group 1.

## Data Availability

Data from systematic review components of the present study are available upon reasonable request to the authors. Data and methods for drug screening will be published separately.

https://camarades.shinyapps.io/relisyr-v1/

## ACKNOWLEDGMENTS

We would like to thank the evidence synthesis and data science communities, including the Collaborative Approach to Meta Analysis and Review of Animal Experimental Studies (CAMARADES) group and developers of R, R Shiny, R Markdown, Tidyverse and SyRF for continued improvement and updates of tools and packages to facilitate the development of SyLECT. We would also like to thank providers of data and resources used in SyLECT, including The Target ALS Human Postmortem Tissue Core, New York Genome Center for Genomics of Neurodegenerative Disease, the Amyotrophic Lateral Sclerosis Association and the Tow Foundation, EMBL-European Bioinformatics Institute.

ReLiSyR-MND consortium members: Charis Wong, Jenna M. Gregory, Jing Liao, Kieren Egan, Hanna M. Vesterinen, Aimal Ahmad Khan, Maarij Anwar, Caitlin Beagan, Fraser Brown, John Cafferkey, Alessandra Cardinali, Jane Yi Chiam, Claire Chiang, Victoria Collins, Joyce Dormido, Elizabeth Elliott, Peter Foley, Yu Cheng Foo, Lily Fulton-Humble, Angus B. Gane, Stella A. Glasmacher, Áine Heffernan, Kiran Jayaprakash, Nimesh Jayasuriya, Amina Kaddouri, Jamie Kiernan, Huw Knapper, Gavin Langlands, Danielle Leighton, Jiaming Liu, James Lyon, Olena Makysm, Arpan R. Mehta, Alyssa Meng, Vivienne Nguyen, Na Hyun Park, Suzanne Quigley, Yousuf Rashid, Andrea Salzinger, Bethany Shiell, Ankur Singh, Tim Soane, Alexandra Thompson, Olaf Tomala, Fergal M. Waldron, Bhuvaneish T. Selvaraj, Jeremy Chataway, Robert Swingler, Peter Connick, Suvankar Pal, Siddharthan Chandran, Malcolm R. Macleod

## CONTRIBUTIONS

Project conceptualisation: CW, AC, SP, SC, MM

Data curation: CW, AC, JLiao, BTS, PB, RC, JLongden

Formal analysis: CW, AC, JLiao, BTS, PB, RC, JLongden, RG, MM

Funding acquisition: SC

Investigation: CW, AC, JLiao, BTS, PB, RC, JLongden, MM, ReLiSyR-MND consortium

Methodology: CW, AC, PB, RC, SP, GH, NC, SC, MM

Project administration: CW, AC, JLiao, BTS

Software: CW, JLiao, AC

Supervision: SP, GH, NC, SC, MM

Validation: CW, AC, BTS, PB, RC

Visualisation: CW, AC, BTS, RG

Drug selection group: CW, AC, BTS, RC, PB, RD, JC, RS, SP, SC, MM

Writing – original draft: CW, AC, BTS

Writing – review and editing: CW, AC, JLiao, BTS, PB, RC, JLongden, RH, RD, SP, JC, RS, GH, NC, SC, MM

## FUNDING

MND-SMART is funded by grants from MND Scotland, My Name’5 Doddie Foundation (DOD/14/15) and specific donations to the Euan MacDonald Centre. SC receives funding from UK Research and Innovation (Medical Research Council) (DRI-CORE-2017-EDI). The study funders have no role in study design, data collection, data analysis, data interpretation or writing of publications. For the purpose of open access, the author has applied a Creative Commons Attribution (CC BY) licence to any Author Accepted Manuscript version arising from this submission.

## COMPETING INTERESTS

In the last 3 years, J. Chataway has received support from the Efficacy and Evaluation (EME) Programme, a Medical Research Council (MRC) and National Institute for Health Research (NIHR) partnership and the Health Technology Assessment (HTA) Programme (NIHR), the UK MS Society, the US National MS Society and the Rosetrees Trust. He is supported in part by the NIHR University College London Hospitals (UCLH) Biomedical Research Centre, London, UK. He has been a local principal investigator for a trial in MS funded by the Canadian MS society. A local principal investigator for commercial trials funded by: Actelion, Novartis and Roche; and has taken part in advisory boards/consultancy for Azadyne, Janssen, Merck, NervGen, Novartis and Roche.

## Notes

### Clinical Protocols

https://osf.io/bkscj

